# Mathematical modeling to inform vaccination strategies and testing approaches for COVID-19 in nursing homes

**DOI:** 10.1101/2021.02.26.21252483

**Authors:** Rebecca Kahn, Inga Holmdahl, Sujan Reddy, John Jernigan, Michael J. Mina, Rachel B. Slayton

**Author notes:** contributed equally (Rebecca Kahn, Inga Holmdahl). The findings and conclusions in this report are those of the authors and do not necessarily represent the official position of the Centers for Disease Control and Prevention (CDC).

## Abstract

**Background:** Nursing home residents and staff were included in the first phase of COVID-19 vaccination in the United States. Because the primary trial endpoint was vaccine efficacy (VE) against symptomatic disease, there are limited data on the extent to which vaccines protect against SARS-CoV-2 infection and the ability to infect others (infectiousness). Assumptions about VE against infection and infectiousness have implications for possible changes to infection prevention guidance for vaccinated populations, including testing strategies.

**Methods:** We use a stochastic agent-based SEIR model of a nursing home to simulate SARS-CoV-2 transmission. We model three scenarios, varying VE against infection, infectiousness, and symptoms, to understand the expected impact of vaccination in nursing homes, increasing staff vaccination coverage, and different screening testing strategies under each scenario.

**Results:** Increasing vaccination coverage in staff decreases total symptomatic cases in each scenario. When there is low VE against infection and infectiousness, increasing staff coverage reduces symptomatic cases among residents. If vaccination only protects against symptoms, but asymptomatic cases remain infectious, increased staff coverage increases symptomatic cases among residents through exposure to asymptomatic but infected staff. High frequency testing is needed to reduce total symptomatic cases if the vaccine has low efficacy against infection and infectiousness, or only protects against symptoms.

**Conclusions:** Encouraging staff vaccination is not only important for protecting staff, but might also reduce symptomatic cases in residents if a vaccine confers at least some protection against infection or infectiousness.

**Summary:** The extent of efficacy of SARS-CoV-2 vaccines against infection, infectiousness, or disease, impacts strategies for vaccination and testing in nursing homes. If vaccines confer some protection against infection or infectiousness, encouraging vaccination in staff may reduce symptomatic cases in residents.

## Background

Nursing homes have been devastated by the ongoing COVID-19 pandemic in the United States [1]. Nursing home residents are disproportionately affected by severe disease and mortality due to their older age and high prevalence of comorbidities. In addition, congregate living and necessarily close contacts (e.g. assistance with activities of daily living) between staff and residents have made controlling outbreaks in these settings more challenging than in the general community. Because of this, residents and staff of long-term care facilities have been included in the Advisory Committee on Immunization Practices’ first phase (1a) for vaccination, alongside healthcare personnel [2]. Vaccine rollout began in nursing homes across the country in late December 2020 and early January 2021.

Screening testing, i.e., testing of asymptomatic individuals, paired with enhanced infection prevention and control (IPC), has been one of the primary strategies available to control outbreaks in nursing homes, although the extent and frequency of testing has been hampered by resource availability [3]. Current recommendations are to test previously undiagnosed residents and staff every 3-7 days if there is an outbreak in the facility. Staff should be tested up to twice weekly regardless of outbreak status, depending on community test positivity [4,5]. While both polymerase chain reaction (PCR) and antigen tests are used, we found in previous work that point of care testing, such as antigen tests – which are less sensitive to detect any virus RNA but nearly as sensitive for infectious virus – may better reduce transmission when used at the same frequency. This is due largely to the rapid turnaround time for results [6,7].

The results of clinical trials of vaccines currently authorized in the United States show high vaccine efficacy (VE) against symptomatic disease across all age groups, which is promising for reducing morbidity and mortality among nursing home residents [8,9]. As of January 2021, over 100,000 residents, representing 19.5% of residents with confirmed infection, and over 1000 staff have died [10]. While mortality rates will vary by age and other factors, the ability of the vaccine to reduce symptomatic disease, especially severe symptoms, will have a substantial impact on mortality.

The vaccine trials did not provide data on VE against all infection (i.e., including asymptomatic infection) or infectiousness (e.g. ability to transmit virus to others, such as by blunting or reducing the duration of peak viral load) [11]. The VE against infection and infectiousness have important implications for understanding whether these vaccines can build herd immunity in a population and for identifying when and to what extent other IPC strategies can be lifted. In nursing homes, outbreak control measures take substantial resources and do have important negative consequences—including restricting visitors and drawing on limited staff time.

Communities of color comprise a large proportion of nursing home staff [12,13]. Recent analyses have found lower vaccination uptake among staff compared to residents [14–16]. This may be due in part to hesitancy stemming from historical injustices that have justifiably resulted in reduced trust in medicine and the safety of a novel vaccine [17].

Here, we use mathematical modeling to examine at the effects of vaccination in nursing homes, with the understanding that vaccination among the elderly in the general community will lag behind vaccination in nursing homes. While vaccinating residents is a priority, we focus here on the effects of increasing vaccination among staff, as high resident turnover makes it challenging to maintain high vaccination levels among residents in the absence of high community vaccination coverage. We also look at testing strategies under different assumptions about the mode of VE to evaluate how, or whether, screening testing recommendations may be changed following vaccine rollout.

## Methods

### Model overview

Here, we expand upon a previously developed stochastic, agent-based Susceptible–Exposed– Infectious (Asymptomatic/Symptomatic)–Recovered (SEIR) model of SARS-CoV-2 transmission in nursing homes [6]. Infected individuals are identified either based on symptoms or through screening testing, after which they are isolated in a separate COVID-19 specific area of the nursing home. Staff in the COVID-19 area are assumed to have access to personal protective equipment. Resident lengths of stay are variable and drawn from a distribution based on data from the publicly available Centers for Medicare and Medicaid Services Minimum Data Set 3.0 from 2016 [18] (Table S1); we made the simplifying assumption that the nursing home remains at 100% capacity, with new residents replacing those who have died or been discharged. We assume new resident admissions are not vaccinated or immune from previous infection, reflecting low community vaccination coverage, but vary this in a sensitivity analysis (Table S1).

The nursing home consists of 100 residents, with two per room, and 100 staff who are assumed to be split evenly across three shifts. Infected staff are not eligible to work, resulting in shortages of staff in the nursing home; if staffing falls below 50%, temporary workers are brought in as replacements. Aside from temporary workers, we assume that there is no staff turnover. We assume residents do not contact other residents, with the exception of their roommates, and daily contact rates between residents and staff are based on contact rates from nursing homes across the United States [19,20].

Viral load is modeled stochastically for each infected individual [6]. Importantly, we assume that viral load trajectories for asymptomatic and symptomatic infections are drawn from the same parameter distributions [21,22]. We model infectiousness categorically, making the assumption that it depends on viral load (i.e. number of RNA copies per mL) as follows: not infectious (<10^4 copies/mL), moderately infectious, i.e., 50% of full infectiousness, and fully infectious (>10^7 copies/mL). While the relationship between infectiousness and viral load is not fully understood, these assumed values fall within the range used in other models [22]. This is likely conservative as peak viral loads routinely exceed 10^10 copies/mL, and full infectiousness may not be reached until viral loads are closer to this range.

### Testing strategies

We evaluate two types of screening tests: 1) rapid antigen testing and 2) PCR testing, simulating either weekly testing or testing every three days (2.3 times per week on average). These testing scenarios are compared to a scenario where testing is only symptom-based, i.e., there is no screening testing of individuals who do not have symptoms. PCR and antigen tests vary in their sensitivity (modeled as viral limit of detection (LOD)) and turnaround time. Based on the data available on these tests, antigen testing has a higher LOD than PCR (Table S1) but returns results immediately, whereas PCR here has a two day delay. For symptomatic individuals, isolation begins immediately upon symptom onset, while for asymptomatic individuals, isolation is not implemented until results are returned. While not explicitly modeled, we assume high specificity of antigen tests would be achieved through rapid confirmatory tests.

### Vaccine efficacy scenarios

We incorporate three types of VE into the model structure: vaccine efficacy against progression to symptoms among those infected (VE_P_), vaccine efficacy against susceptibility to infection (VE_S_), and vaccine efficacy against infectiousness (VE_I_) among those infected (Table 1) [11]. In all simulations, we assume vaccine efficacy against symptoms - a combination of VE_S_ and VE_P_ - is 90%, which is similar to the findings from the Pfizer and Moderna vaccine trials [8,9]. Because these trials only provided data on vaccine efficacy against disease (i.e. symptomatic infection), we compare three different scenarios (Table 1) that would all result in a total VE against symptoms of 90% but vary in their efficacy against susceptibility to infection and infectiousness. In the first scenario, we assume that VE against symptoms comes entirely from efficacy against susceptibility to infection (VE_S_ = 90%) and that efficacy against infectiousness is also high. In the second scenario, we assume a lower efficacy against susceptibility to infection and infectiousness and therefore a higher VE_P_ than in the first scenario. In the final scenario, we assume the only protection the vaccine confers is against progression to symptoms (VE_P_ = 90%). In all three scenarios, partial efficacy is conferred seven days after the first dose, and full protection is conferred seven days after the second dose. Doses are administered 21 days apart.

**Table 1.** Parametrization of vaccine efficacy (VE) scenarios with varying levels of protection against infection and infectiousness **

|  | VE parameters for given scenario |  |  |
| --- | --- | --- | --- |
| VE Scenario | Vaccine Efficacy against Progression to Symptoms (VE<sub>P</sub>)<sup>*</sup> | Vaccine Efficacy against Susceptibility to Infection (VE<sub>S</sub>)<sup>*</sup> | Vaccine Efficacy against Infectiousness (VE<sub>I</sub>)<sup>*</sup> |
| 1: VE against Symptoms, Infection, Infectiousness (High) | 0%, 0% | 50%, 90% | 50%, 90% |
| 2: VE against Symptoms, Infection, Infectiousness, (Low) | 33%, 80% | 25%, 50% | 25%, 50% |
| 3: VE against Symptoms only | 50%, 90% | 0%, 0% | 0%, 0% |

<sup>\*</sup> First value is the efficacy of the first dose, and second value is the efficacy after the second dose.
<sup>\*\*</sup> To reflect results of vaccine trials, each scenario has a final vaccine efficacy against symptoms of 90% after two doses, which is a combination of VE<sub>S</sub> and VE<sub>P</sub>

To isolate the effects of the different vaccine scenarios in our analysis, everyone in the facility is susceptible to infection at the beginning of each simulation and no cases are introduced into the facility until 8 days after the second dose, after which staff have a daily probability of infection from the community. In a sensitivity analysis, we examine the impact of allowing cases to be introduced 8 days after the first dose. To identify the effects of vaccination among staff, 90% of residents are vaccinated with two doses at baseline, and we only vary coverage among staff. Our primary endpoint for each of these vaccine scenarios and testing strategies is cumulative symptomatic infections after 40 days, using the mean across 100 simulations.

## Results

As expected, we find that in each of the VE scenarios, increasing vaccination coverage among staff reduces total symptomatic cases (i.e. among both residents and staff) (Figure 1). The scenario with high VE against infection and infectiousness (Scenario 1) is most effective at reducing total symptomatic cases. When the vaccine only protects against symptoms (Scenario 3), total symptomatic cases are highest within each level of staff vaccination coverage.

**Figure 1.**
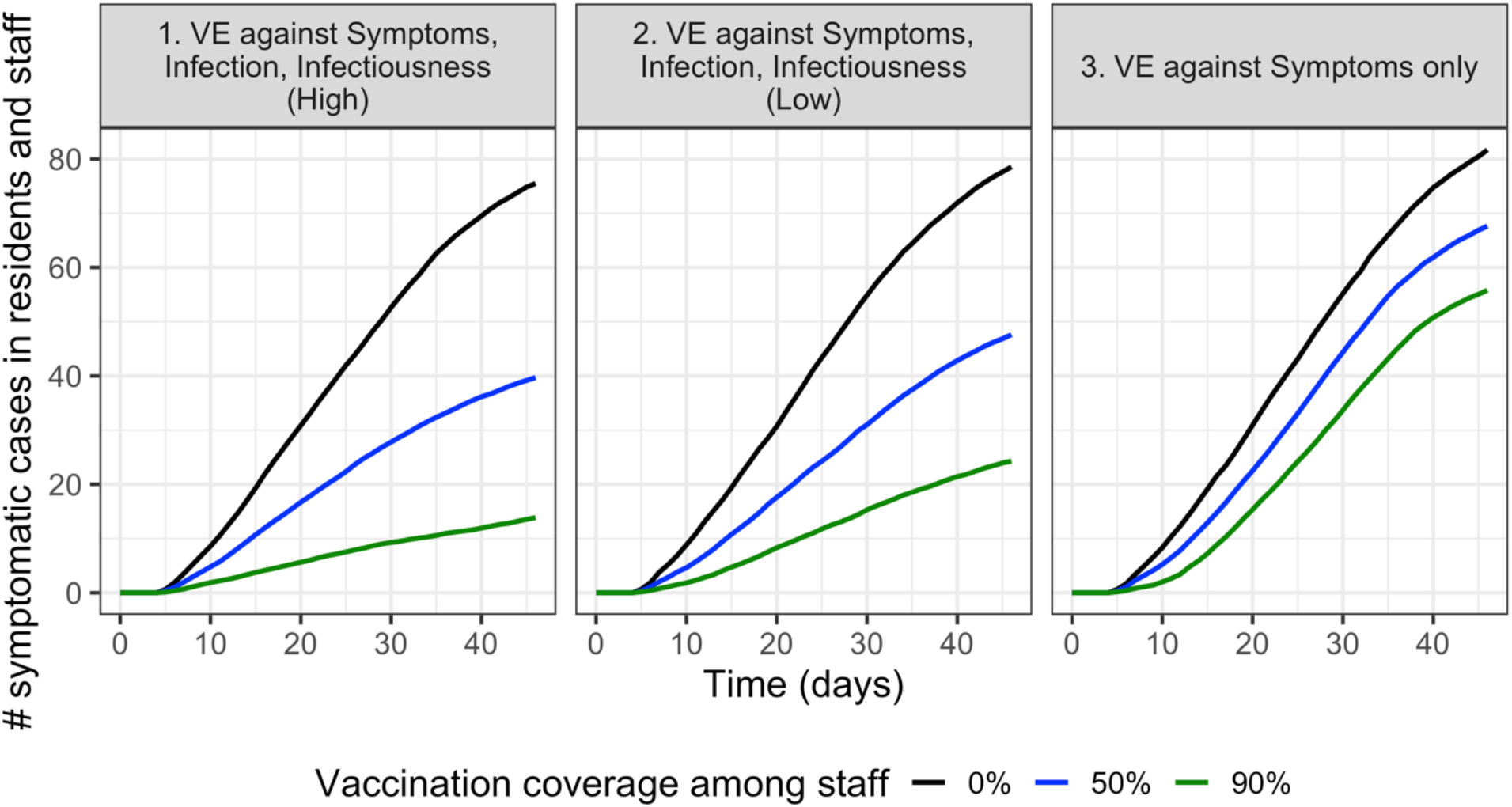
In each of the vaccine efficacy scenarios, increasing vaccination coverage among staff reduces total symptomatic cases (i.e. combined symptomatic cases among residents and staff). Two dose vaccinations are assumed to be completed before cases are able to be introduced into the facility starting on day 0.

While increasing vaccination coverage among staff reduces overall symptomatic cases across both staff and residents in all three scenarios, the impact of staff vaccination coverage on total symptomatic cases in *residents only* is highly dependent on the type of vaccine efficacy (Figure 2). When the vaccine protects against infection and infectiousness, increasing vaccination coverage among staff reduces symptomatic cases among residents. The importance of staff vaccination to protect resident infections in these scenarios is likely due to low coverage among residents. Although initial vaccine coverage is 90% in residents, there is high resident turnover and low assumed community vaccination coverage, so the proportion of residents who are vaccinated falls quickly. In a sensitivity analysis where 50% of incoming residents are assumed to be vaccinated, the total number of symptomatic cases is lower, but staff coverage still remains important for reducing cases among residents, and we see the same trends across vaccine efficacy scenarios (Figures S1).

**Figure 2.**
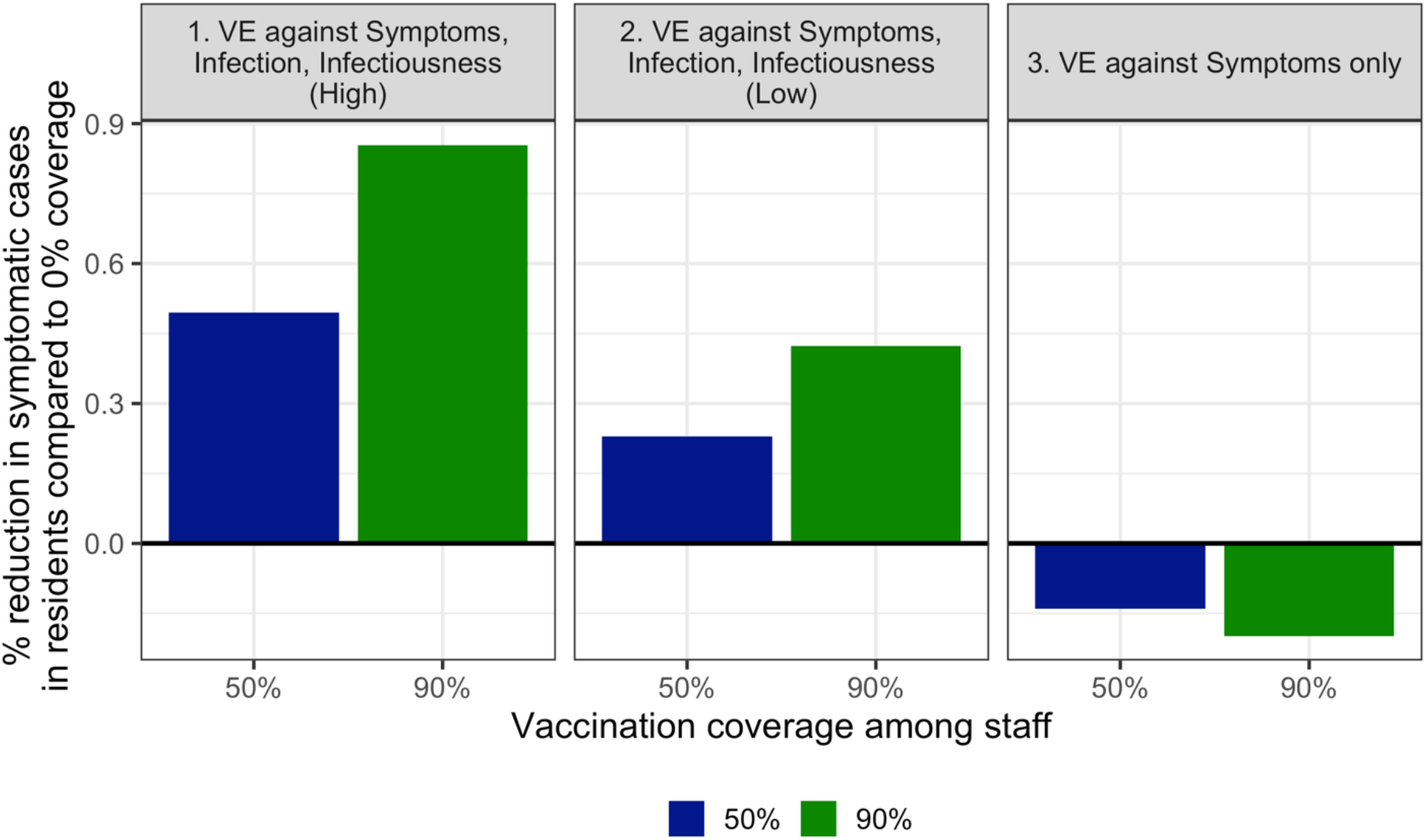
When considering symptomatic cases in residents, increasing vaccination coverage among staff from 0% reduces cases in vaccine scenarios 1 and 2, when vaccination confers at least low protection against infection and infectiousness. When the vaccine protects against symptoms only, increasing vaccination coverage among staff increases symptomatic cases among residents.

On the other hand, if the vaccine only protects against symptoms, higher coverage among staff may increase the proportion of cases that are asymptomatic, leading to more undetected transmission. This leads to higher total symptomatic cases among residents as staff coverage increases (Figure 2, panel 3).

The importance of continued screening testing also varies by type of vaccine efficacy (Figure 3). When the vaccine has partial or no efficacy against infections and infectiousness (Scenarios 2 and 3), more frequent screening testing reduces total symptomatic cases in residents. When VE protects against symptoms only, the vaccine increases the proportion of asymptomatic infections, while also failing to prevent infections and induce herd immunity; therefore, high frequency of screening testing is particularly important for controlling disease spread. Due to faster turnaround time, antigen testing results in lower incidence than PCR testing done at the same frequency. If the vaccine has high efficacy against infection and infectiousness (Scenario 1), screening testing has little added benefit over symptom-based testing. In a sensitivity analysis where cases are allowed to be introduced 8 days after the first dose (compared to 8 days after the second dose in our baseline scenario), we see that screening testing is important for reducing outbreak size under all VE scenarios (Figure S4).

**Figure 3.**
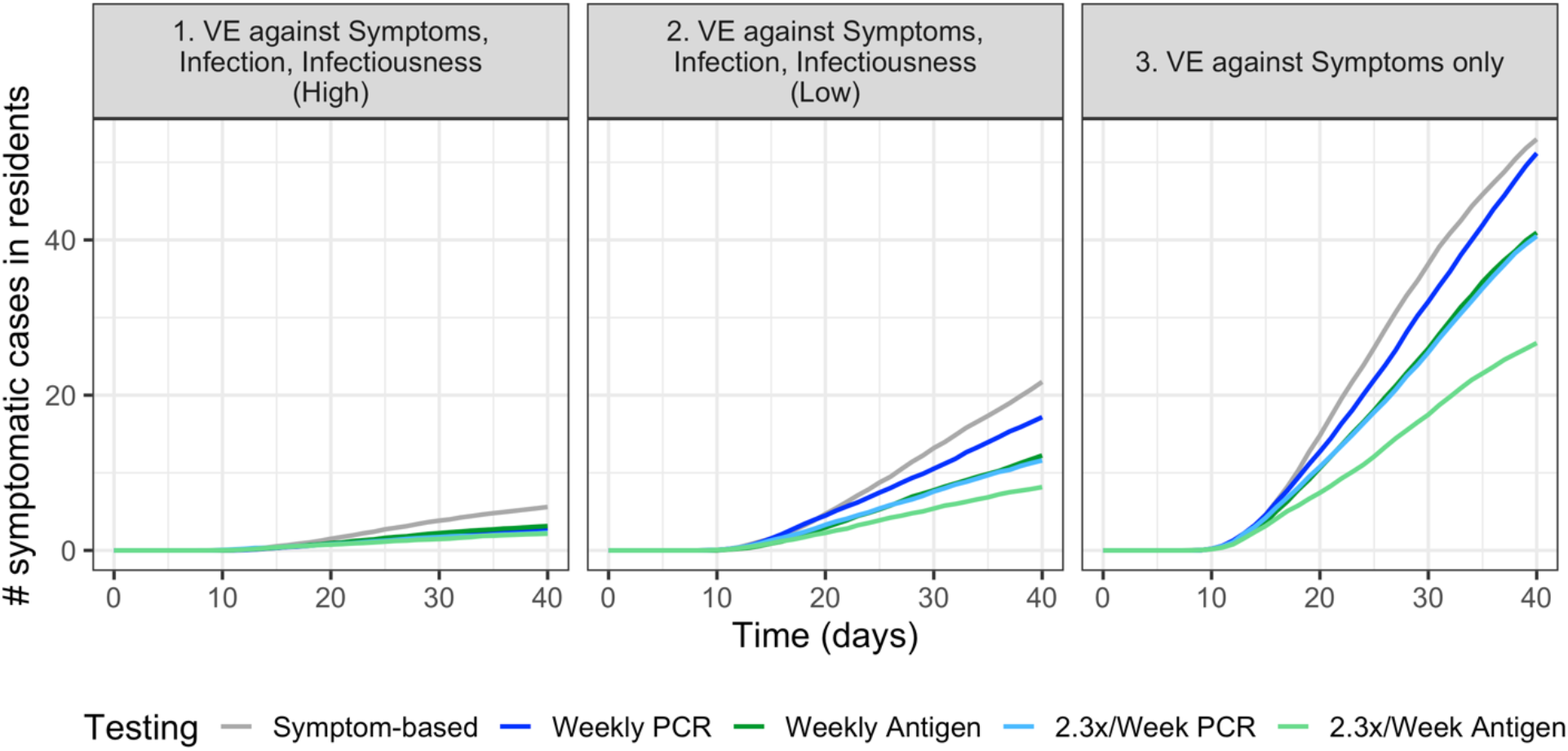
The differences between testing strategies vary across vaccine efficacy scenarios. When the vaccine has low or no efficacy against infections and infectiousness (Scenarios 2 and 3), frequent screening testing is important for reducing total symptomatic cases in residents. Due to faster turnaround time, antigen testing results in lower incidence than PCR testing at the same frequency.

## Discussion

As vaccine program implementation continues in nursing homes and across the country, understanding how well vaccines are able to reduce infection and infectiousness in addition to symptomatic infection is critical for informing ongoing strategies to control COVID-19 outbreaks. By modeling outbreaks within a single nursing home, we look at the impact of vaccination coverage among staff and multiple testing strategies under different assumptions about vaccine efficacy. We find that given high resident turnover in nursing home settings, staff vaccination coverage is a critical factor driving outbreak size. Due to limited vaccine supply, vaccine program implementation will likely continue to be targeted based on risk for some period; in this paper we focus on this period in which vaccines are available but are not yet being offered to the community at large.

We find that increasing vaccination coverage among staff can have a protective effect for residents if the vaccine provides at least some protection against infection and infectiousness. These results highlight the importance of encouraging vaccination among staff—both for their protection, as they have been one of the groups most severely affected to date, and also to protect residents, who are among those at highest risk of mortality. Efforts to increase staff vaccination should include culturally competent messaging and support to address the concerns of nursing home staff. Reaching higher vaccine coverage among staff could allow for less screening testing, as we see that increased frequency of testing has little added benefit when efficacy against infection is high.

If the vaccine does not protect against infection and infectiousness, but only against symptoms, our analysis indicates that increasing staff coverage might lead to higher numbers of symptomatic cases among residents. These results underscore the importance of continuing to provide frequent screening testing in nursing homes, particularly during outbreaks, until more data are available on the types and extent of protection the vaccines provide. Given the importance of rapid results, we see that point of care tests, such as antigen tests, may be more effective than PCR tests in reducing outbreak size, particularly in areas that do not have access to rapid PCR test results. Newer high sensitivity PCR-quality rapid tests like rapid loop-mediated isothermal amplification (LAMP) assays may become available soon, which would provide speed and accessibility without compromising sensitivity.

An important driver of the results from this model is relatively low vaccine coverage among residents, even after high coverage during initial vaccine rollout, due to the assumptions of high resident turnover and low community vaccination rates. Once community vaccination coverage is high enough that most incoming residents are already protected, the impact of staff coverage and the importance of testing symptomatic cases in residents will diminish. However, due to the timing and challenges with vaccine rollout so far in many areas and low seroprevalence in older age groups [23], reaching these levels will likely take many months.

We have made simplifying assumptions about the logistics of vaccine rollout, with only two days for vaccination (one for each dose). Many nursing homes conducted vaccinations across three days, and in the future residents may be offered vaccination upon admission to the nursing homes, which would help maintain high levels of resident vaccination coverage in the short-term. Given the two-dose schedule, many residents will not stay long enough to see the effects of both doses, or even get a second dose, even if opportunities for vaccination are provided during their stay (Table S1). This underscores the importance of continuing to vaccinate community-dwelling individuals (e.g., older adults, those with multiple comorbidities) before they become nursing home residents to ensure that short-stay residents are protected. We have also made several assumptions regarding contact patterns, including modeling staff as one population; in reality, nursing homes employ many different types of staff who have different responsibilities and levels of interaction with residents that affect risk of transmission. We made the conservative assumption of equal infectiousness given vaccination status for asymptomatic and symptomatic infections. We also did not specifically assess the potential benefits of outbreak vs non-outbreak testing in a vaccinated population, which may have important implications for prioritizing limited testing resources; we focus on screening testing in this analysis. We further assume nursing homes will maintain strict policies limiting visitation and socializing within the facility over the simulation’s time horizon. These policies have important implications for quality of life and have been one of the major challenges for nursing home residents during the pandemic. Future research is needed to explore the impact of vaccine efficacy, testing practices, and community incidence rates on the ability to safely relax these and other IPC policies.

Data from vaccine rollout in nursing homes and other settings prioritized for early vaccination have the potential to improve our understanding of the mode(s) and level of vaccine efficacy. These data may also provide insight into the efficacy of these vaccines against new variants of concern. Until there is sufficient evidence indicating the extent of efficacy against infection or infectiousness, screening testing remains a key tool for mitigating outbreaks in these high-risk settings, and frequent testing should continue to be conducted to prevent nursing home outbreaks [24].

## Data Availability

Code is available on github.

https://github.com/rek160/NursingHomeVaccineModel

## Acknowledgements

We thank Matt Samore for helpful discussion.

## Supplementary Materials

**Figure S1.**
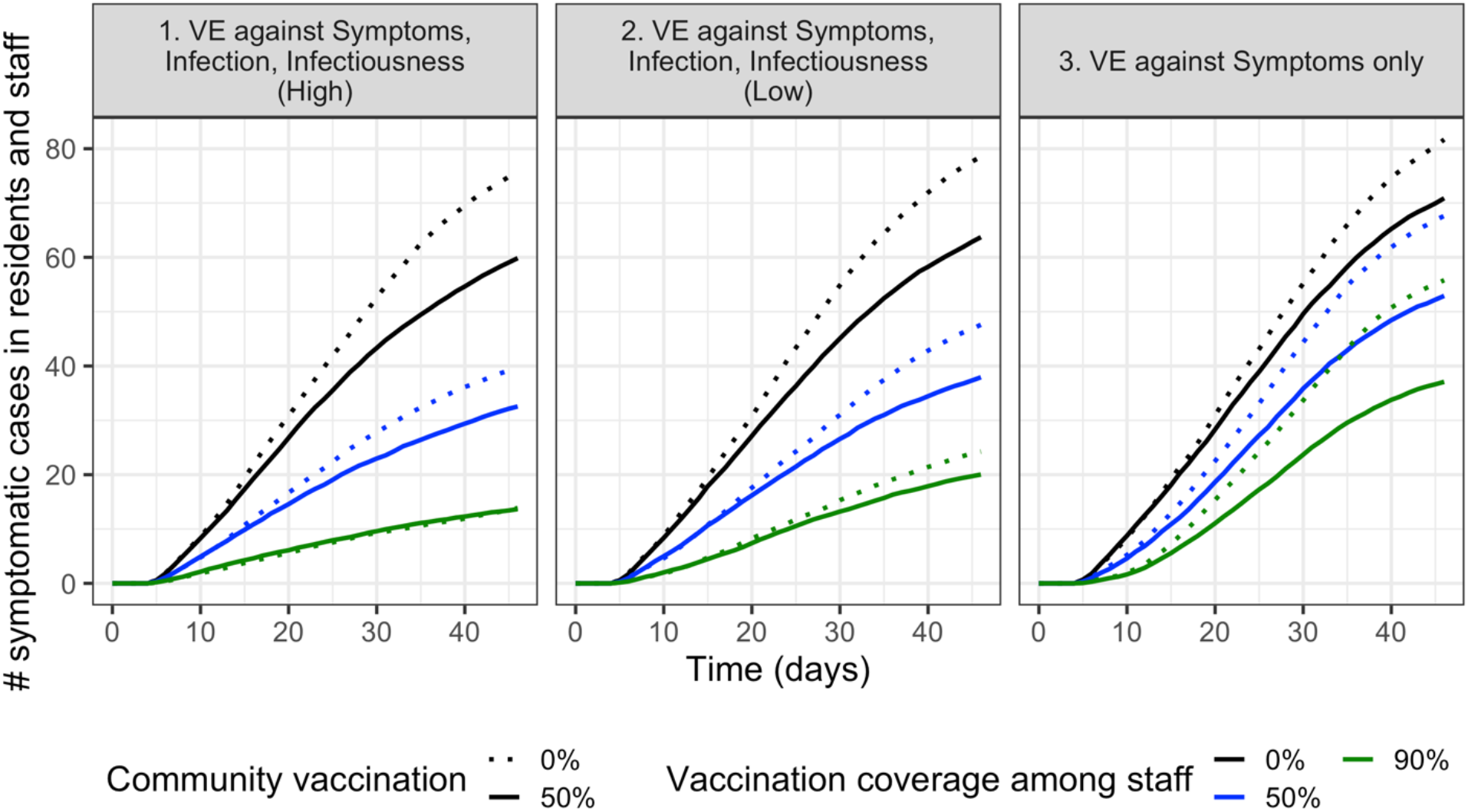
Dotted lines show original simulations from Figure 1 with no vaccination in the community. Solid lines show means of simulations done with 50% of incoming residents already vaccinated.

**Figure S2.**
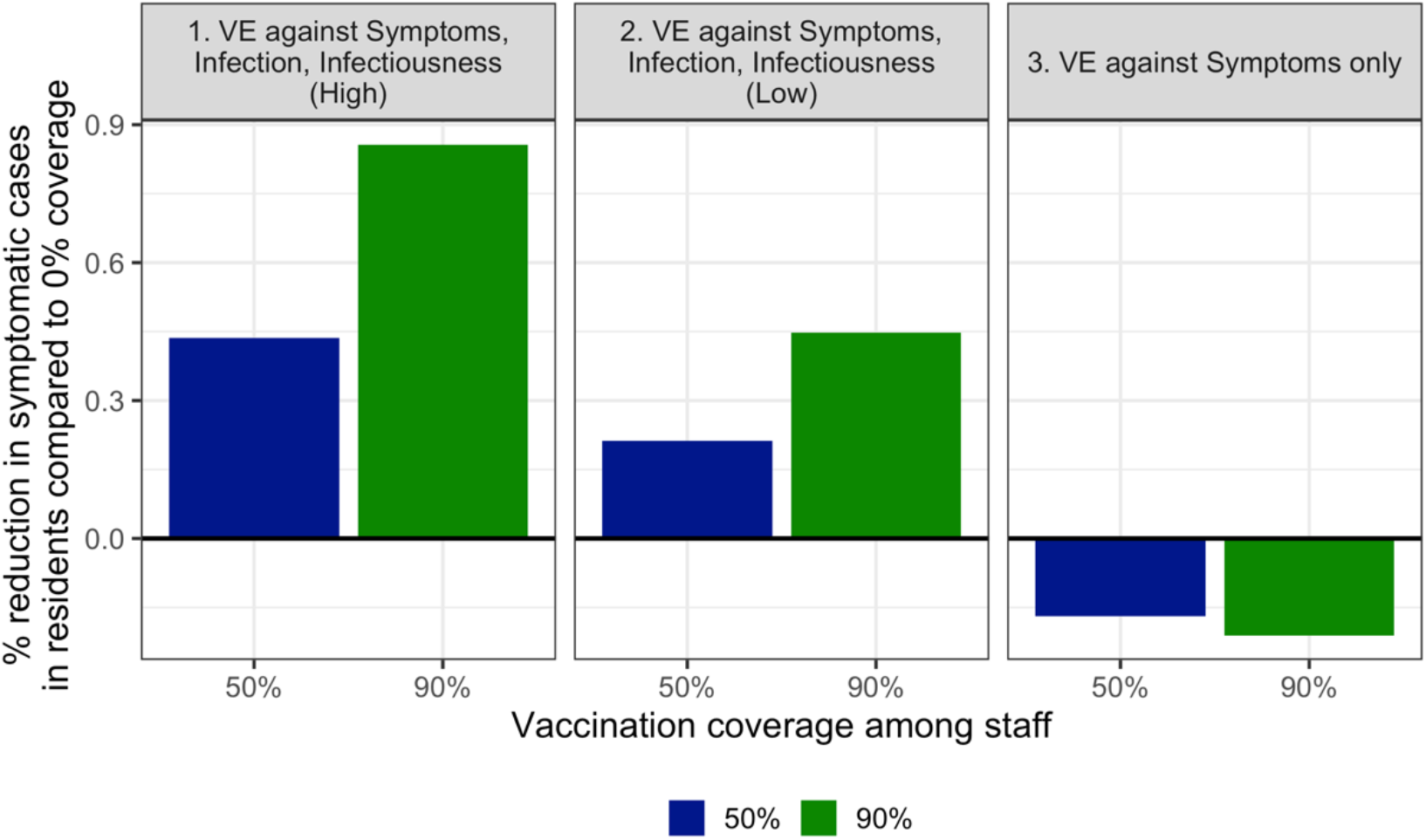
Effect of staff vaccination when 50% of incoming residents are vaccinated. When considering symptomatic cases in residents, increasing vaccination coverage among staff from 0% reduces cases in vaccine scenarios 1 and 2, when vaccination confers at least low protection against infection and infectiousness. When the vaccine protects against symptoms only, increasing vaccination coverage among staff increases symptomatic cases among residents.

**Figure S3.**
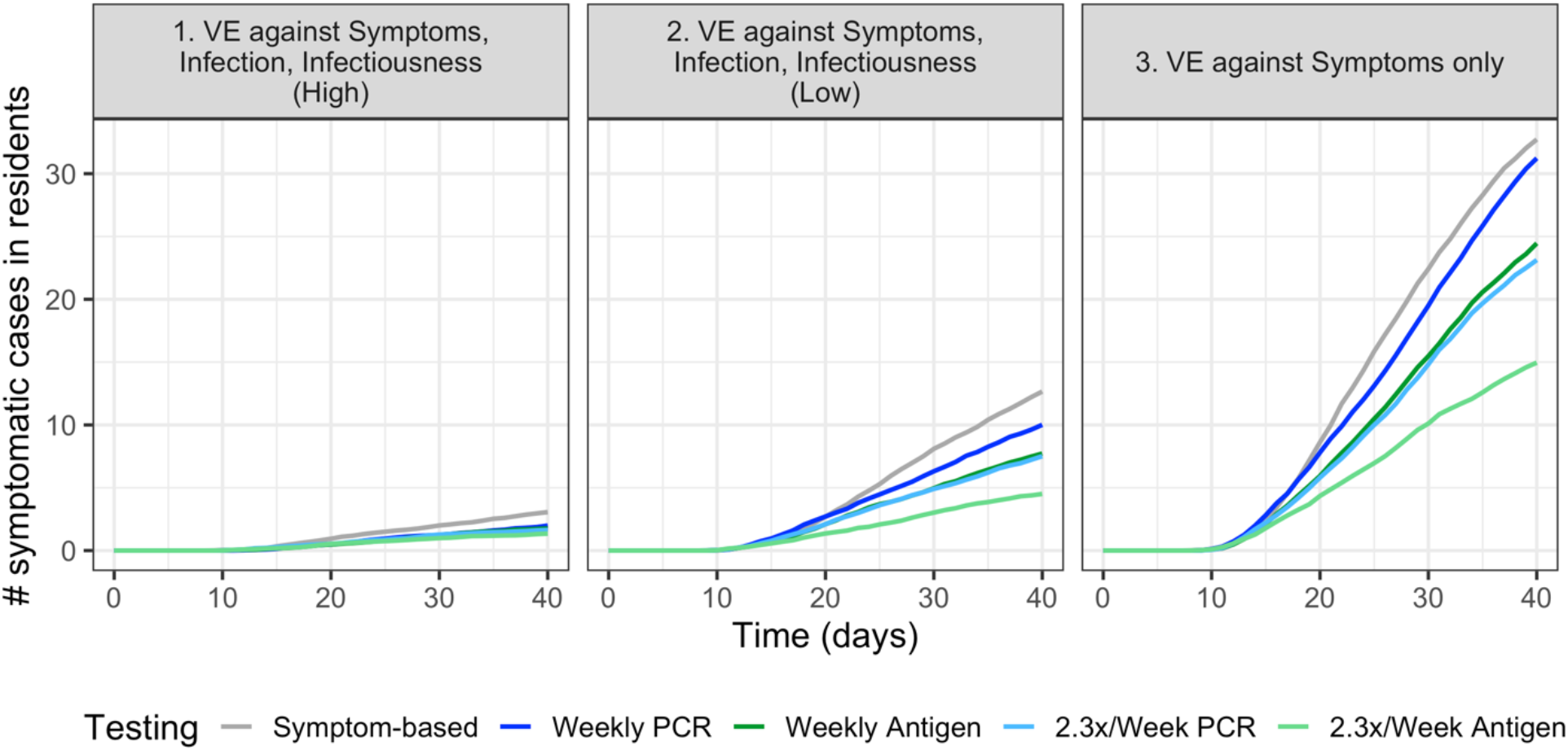
Effect of testing when 50% of incoming residents are vaccinated. When the vaccine has low or no efficacy against infections and infectiousness (Scenarios 2 and 3), frequent screening testing is important for reducing total symptomatic cases in residents. Due to faster turnaround time, antigen testing results in lower incidence than PCR testing at the same frequency.

**Figure S4.**
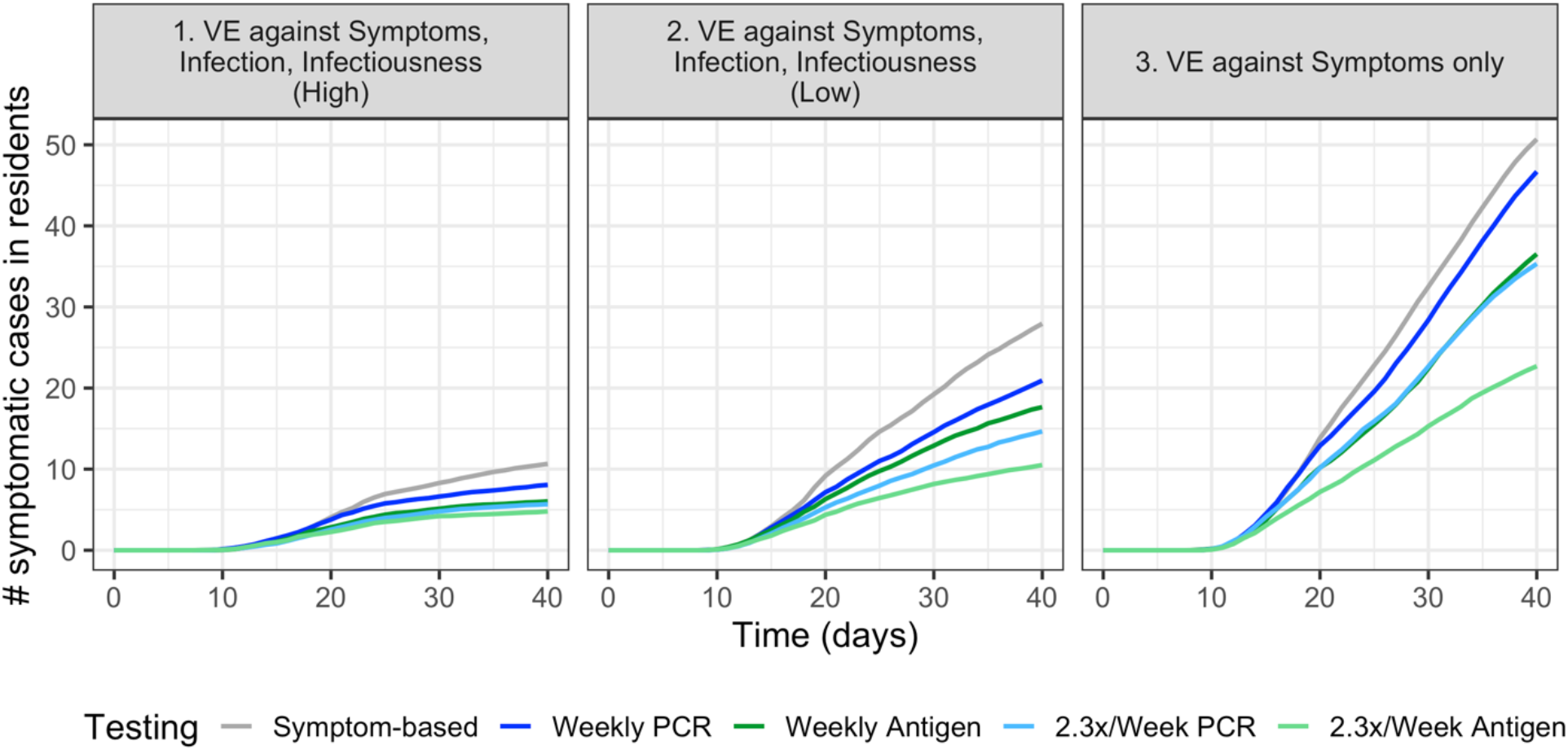
When cases are allowed to be introduced 8 days after the first vaccine dose (vs. 8 days after the second dose in Figure 3), we see that testing remains more important for controlling outbreak size under all VE scenarios. Here, the second dose is administered on day 13, and takes effect on day 20.

**Table S1.** Model parameters

| Parameter | Values* |
| --- | --- |
| Number of residents | 100 [19] |
| Number of staff | 100 [19] |
| Probability of infection per infectious contact | 0.02 |
| Latent period (days) | 3-5 [25] |
| Time in infectious compartment (days); infectiousness dependent on viral load | 14 [6] |
| Daily probability of infection from the community | 0.03 |
| Daily contacts staff-staff | 2 [19] |
| Daily contacts residents - staff | 6 [19,20] |
| Daily contacts staff - residents | 6 - 12 [19] |
| Daily contacts residents - residents (non-roommates) | 0 [19] |
| Proportion of staff asymptomatic | 0.4 [26,27] |
| Proportion of residents asymptomatic | 0.2 [28–30] |
| Duration of presymptomatic transmission (days) | 2 [25,27,31] |
| Reduction in force of infection per contact from PPE | 95% [32] |
| Proportion of temporary staff previously infected (and assumed immune) upon entry into nursing home | 0.2 |
| Proportion of incoming residents vaccinated | 0, 50% |
| Baseline mortality (daily) | 1/1000 |
| COVID-19 mortality (daily) | 2/100 |
| Mean peak viral load (copies/mL) | 10<sup>8</sup> [33] |
| Limit of detection - rapid antigen test (copies/mL) | 10<sup>5</sup> [22, 34–36] |
| Limit of detection - PCR (copies/mL) | 10<sup>3</sup> [22, 37] |
| Antigen and PCR test specificity | 1 |
| Viral load threshold for infectiousness (copies/mL) | $10^4$ |
| Viral load threshold for high infectiousness (copies/mL) | $10^7$ |
| Turnaround time - antigen test (days) | Same day |
| Turnaround time - PCR test (days) | 2 |
| Time between vaccine dose 1 and 2 (days) | 21 [9] |
| Time until effect of vaccine dose after vaccination (days) | 7 |
| Length of stay 25<sup>th</sup>, 50<sup>th</sup>, 75<sup>th</sup> percentiles, after restricting to minimum of 7 days (days) | 15, 27, 66 [18,38] |

\*If no reference is cited, parameters are assumed.

